# Nanopore-based pathogen surveillance allows complete metagenome-assembled genome reconstruction of low-abundance enteric pathogens in wastewater samples

**DOI:** 10.1101/2025.03.19.25324250

**Authors:** Jeff Gauthier, Sima Mohammadi, Irena Kukavica-Ibrulj, Brian Boyle, Chrystal Landgraff, Lawrence Goodridge, Roger C. Levesque

## Abstract

**Background:** Pathogen surveillance often relies on culture-based methods and epidemiological traceback investigations that are both time- and cost-ineffective, especially in the case of enteropathogenic bacteria contaminating food supplies. Nevertheless, metagenomic sequencing of wastewater influent helps conduct continuous, culture-independent, and community-level monitoring of microbes shed from the human gut microbiome. However, even though amplicon-based microbiome census methods help identify taxa, they typically do not allow strain-level epidemiology or investigating virulence factors and antimicrobial resistance mechanisms involved in an outbreak. Conversely, whole metagenome shotgun sequencing allows both taxonomic surveys and genome reconstruction.

**Results:** Here we present a metagenomic “tracking and assembling” workflow, applied between September 2023 to January 2024, in which we tracked two low-abundance enteric pathogens (Shiga toxin-producing *Escherichia coli* and enteropathogenic non-typhoidal *Salmonella enterica;* 0.1-1% total reads) and reconstructed 95-99% complete genomes using a combined taxonomic read binning and reference-based assembly. Furthermore, for these two pathogens, a maximum abundance peak significantly above baseline levels, assuming 95% confidence, was detected and found to precede by a month two public food recalls, all within the same urban community where municipal wastewater sampling was conducted (Quebec City, Canada).

**Conclusions:** This present work suggests that a continuous “tracking and assembling” approach enhances the resolution of low-abundance pathogen monitoring to the strain level, while also providing information about the gene contents of low-abundance enteropathogens, even when relative abundance is too low to reconstruct genomes via a generic *de novo* assembly and contig binning approach.

## Introduction

Pathogen surveillance has proven critical, especially amid the COVID-19 pandemic, to prevent and control outbreaks. However, conventional monitoring methods often rely on latent indicators, such as case reports from hospitals and samples from infected individuals to both identify the causative agent and its prevalence. This process requires days, even weeks, before mitigating efforts can be deployed.

In parallel, the growing accessibility of high throughput, long read sequencing technology allows to constantly sample a microbiological reservoir and monitor its microbial components to identify emerging threats in a culture-independent manner. To this regard, wastewater is an excellent biological reservoir as it gives a community-level overview of microbial pathogens shedding in human excreta [1].

One area of concern is not only the epidemiology of pathogens themselves, but the one of antimicrobial resistance (AMR). Even though an annual casualty rate of 10 million was predicted by 2050 [2], AMR could have already reached the stage of 1-2 million excess deaths per year as of now [3]. There is a need to identify the causative agents of outbreaks and how they could evade (or gain resistance to) current treatment options.

High throughput sequencing of taxonomic marker genes (e.g. 16S rRNA gene) is routinely used in microbiome surveys [4] but provides insufficient resolution to identify pathogens at the strain level [5] unless long reads are used to sequence the full-length gene [4]. Moreover, marker gene sequencing provides no information about genes of interest that the surveyed taxa carry. Even though functional prediction algorithms from amplicon data have been developed, they lack the sensitivity to indicate biologically meaningful changes between samples [6].

Conversely, whole metagenome shotgun sequencing allows the sampling of all genetic material in a sample [7]. Given enough sequencing depth, metagenome-assembled genomes can be reconstructed [8, 9]. For instance, MaxBin and MetaBAT2 group contigs having similar G-C content, depth of coverage, and tetranucleotide usage, assuming that contigs should share these properties if belonging to the same source organism [8, 9]. However, difficulties arise when co-occurring organisms share these properties, or when the organism of interest is in low abundance among a large set of reads [10].

In a two-year metagenomic survey of Quebec City’s wastewater treatment influent, we compiled taxonomic profiles across weekly samples; however, our targets of interest were primarily enteric pathogenic bacteria whose abundance was often below 1% of total reads. Consequently, reconstructing MAGs for those taxa of interest proved difficult, as the bins often belonged to higher-abundance taxa beyond the scope of our survey. Furthermore, most *de novo* binning algorithms assume that all sequence elements within a genome share the same nucleotide usage and coverage properties, which might not be the case for horizontally exchanged elements (plasmids, prophages) and genomic islands. Furthermore, high-copy-number plasmids may also be excluded from bins because of their higher coverage relative to baseline depth.

We therefore reasoned that sub-setting reads assigned to the species level, and reference-based alignment and assembly, could help recover strain-level assemblies of pathogens of interest. We proceeded as such with Oxford Nanopore reads linked to a common abundance peak for *Escherichia coli* and *Salmonella enterica*. This abundance peak (first detected September 13^th^, 2023) was ultimately found among a subset of 17 samples collected between from September 2023 to January 2024. The pre-assembly read selection ultimately increased genome completeness and reduced contamination while also improving taxonomic assignment compared to a simple reference-based assembly without sub-setting reads. We were also able to capture key virulence factor genes in the resulting assemblies (*stxAB* for STEC, type three secretion system for *S. enterica* serovar *Typhimurium*., further confirming the presence of STEC and virulent non-typhoidal *Salmonella* in this abundance peak.

## Methods

### Wastewater sampling procedure

From September 2023 to January 2024, weekly samples of 63 mL raw sewage water (SW) were collected from the Quebec City Wastewater Treatment East Station, which covers a population of 300,000 including 4 hospitals (**Figure 1**). Immediately upon arrival, total bacterial biomass was enriched by treatment with 600 µL NanoTrap Microbiome B magnetic beads per 50 mL SW (Ceres Nanosciences, Inc.).

**Figure 1.**
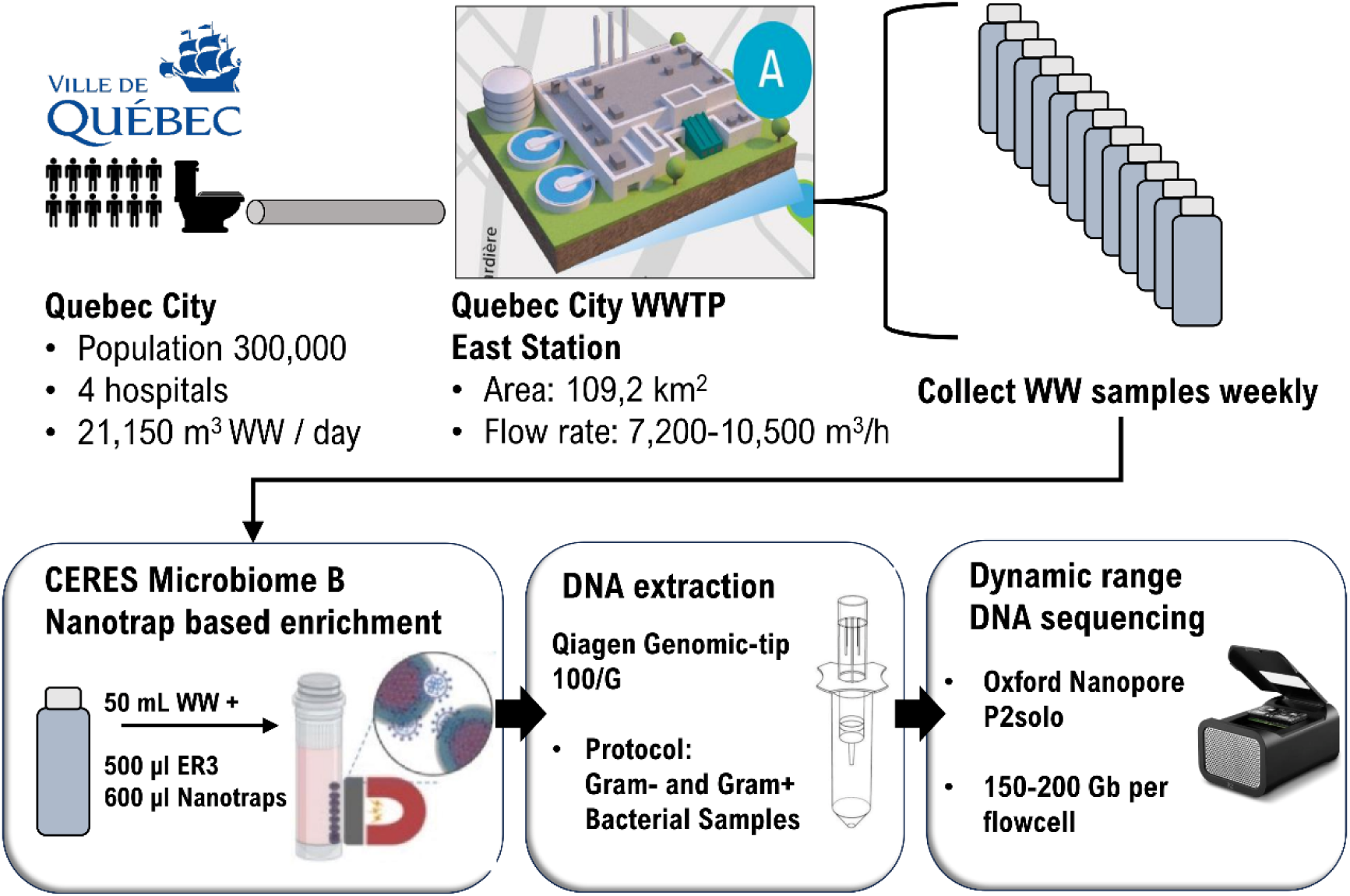
Wastewater sample collection and processing workflow prior to whole metagenome nanopore sequencing. WW: raw wastewater. WWTP: wastewater treatment plant. P2solo: Oxford Nanopore PromethION 2 solo portable DNA sequencer.

### DNA extraction and quantification

Genomic DNA was extracted from cell pellets using the QIAGEN Genomic Tip 100/G kit with the manufacturer’s recommended protocol for Gram-negative and Gram-positive bacteria. DNA extracts were then size selected with the PacBio Short Read Eliminator XS magnetic beads, which completely excludes fragments below 5kb to favor sequencing longer fragments. Size distribution was assessed before and after SREXS treatments with the FemtoPulse automated PFGE platform (Agilent Technologies). Final processed extracts were quantified with the Qubit dsDNA BR method (Thermo Fisher).

### Oxford Nanopore long read sequencing

Throughout the monitoring timeline, samples were sequenced using Oxford Nanopore Technologies (ONT)’s PromethION 2 solo platform. Briefly, 1 µg input DNA was processed with the Ligation Sequencing Kit v14 (SQK-LSK114) as recommended by the manufacturer’s protocol with a few minor adjustments (see Supplementary Methods). Prepared libraries were then sequenced on FLO-PRO002 (P2solo) and basecalled in real-time with MinKNOW v22.04, ONT’s proprietary firmware for sequencing. Base calling was performed using a minimum read length of 300 nt and a quality filter and Q9.

### Taxonomic assignment and baseline monitoring

Reads from the SENTINEL time points were classified with Kraken2 v2.1.2 [11], using the pre-built NCBI Nucleotide k-mer index (761 GB, version 2023-05-22) built by Langmead et al. (https://benlangmead.github.io/aws-indexes/k2). Kraken2 reports were concatenated with the addition of a sample name column for further analysis with R [12] and RStudio [13].

By default, Kraken2 reports percent abundances by the total number of reads per time point. By doing so, reads that could not be classified due to poor quality or length remain included in the calculation. To circumvent this, percent abundances were recalculated over reads that could be classified minimally to the domain level. We postulate that reads of sufficient quality should at least be recognizable as belonging to the known tree of life.

In addition, Kraken2 does not consider abundance cutoffs to report taxa, meaning that one spurious taxon from one single read could be reported. To avoid considering false positives, every count below 100 reads was discarded. As a further sanity check, the abundances should fluctuate over the time series, i.e. taxa should not appear and disappear completely. Therefore, taxa that were reported in less than 75% of the data points were discarded.

### *De novo* metagenome assembly

Nanopore reads from the SW2023-09-13 sample were quality filtered with chopper v0.7.0 (https://github.com/wdecoster/chopper) with minimum Phred score of 12 and minimum read length of 4000 bp. Then, sequencing adapters were trimmed with PoreChop-ABI v0.5.0 [14]. Pre-processed reads were then assembled *de novo* with Flye v2.9.5 [15] with the “—meta” flag to account for uneven contig coverage given the metagenomic nature of the sample. Finally, the draft assembly was polished with Medaka v2.0.1 (https://github.com/nanoporetech/medaka) using pre-processed reads as input bases.

### Reference-guided genome reconstruction

If a known pathogen is present at sufficiently high levels in a SW sample, then it should be possible to reconstruct its genome sequence, or an approximate consensus of it, using a reference-guided assembly. We therefore pre-selected reads that were identified by Kraken2 as belonging to the same species. Then, only those reads were used for assembly. Briefly, seqtk (https://github.com/lh3/seqtk) was used to subset reads, in conjunction with grep over the Kraken2 reports to extract the list of read IDs. Then, those reads were aligned to a reference using minimap v2.28 [16] using profile “map-ont”. RefSeq genome accessions GCF_000008865.2 and GCF_000006945.2 were used for STEC and nontyphoid *Salmonella enterica* respectively. After alignment, a draft consensus sequence was generated with racon v1.5.0 [17]. A final polishing step was done with medaka v1.11.3 (https://github.com/nanoporetech/medaka) to correct errors intrinsic to Nanopore sequencing. The resulting sequences were evaluated with CheckM v1.2.2 [18] to assess genome completeness in comparison with the reference used. Genome maps were produced with the ProkSee web annotation server [19]. Classical MLST analysis was performed on reconstructed genomes along with their respective references with PyMLST v2.1.6 [20] using the “claMLST command” and the PubMLST datasets for *Salmonella enterica* and *Escherichia coli* (https://pubmlst.org/data/).

### Virulence factor assessment

Genomes (references and draft assemblies) were both re-annotated with Prokka v1.14.3 [21]. Protein FASTA files from Prokka were then given as input to the “VFanalyzer” module of the Virulence Factor Database [22] to generate a presence-absence matrix for known virulence factor genes in *E. coli* and *S. enterica* respectively. In the case of Shiga toxin subunit genes, a pairwise alignment between the Prokka annotation and the reference DNA sequence was done with FastA version 36.3.8 (https://github.com/wrpearson/fasta36).

## Results and Discussion

### Taxonomic assignment and baseline monitoring

From September 2023 to January 2024, 2,796 species were assigned to metagenomic reads throughout the time series (**Suppl. Table 1**). From those, *E. coli* accounted for 0.34% – 0.46% of all classified reads while *S. enterica* accounted for 0.03 – 0.04% of all classified reads (95% confidence interval, N=18), making them 32^nd^ and 245^th^ in rank, respectively. Interestingly, both species reached a maximum abundance peak on September 13^th^, 2023 significantly above the upper confidence limit (0.60% and 0.05% respectively), indicating a surge in bacterial load above baseline fluctuations (**Figure 2** and **Figure 3**). This peak, at the summer-fall transition, is also in agreement with previous studies that reported higher STEC and *Salmonella* cases in the food production chain during this period [23–25]. Of special interest, concerning *Salmonella*, the maximum abundance peak also coincides with two public health notices emitted by Canada’s health regulatory agency, which respectively reported surges of salmonellosis linked to cantaloupes, raw pet food and cattle in six provinces, including Quebec where this study took place [26, 27]. Both notices were first emitted in October 2023, one month after the detected abundance peak. This indicates not only the concordance with clinical/epidemiological observations, but also reinforces metagenomic monitoring as an early indicator of potential outbreaks.

**Figure 2.**
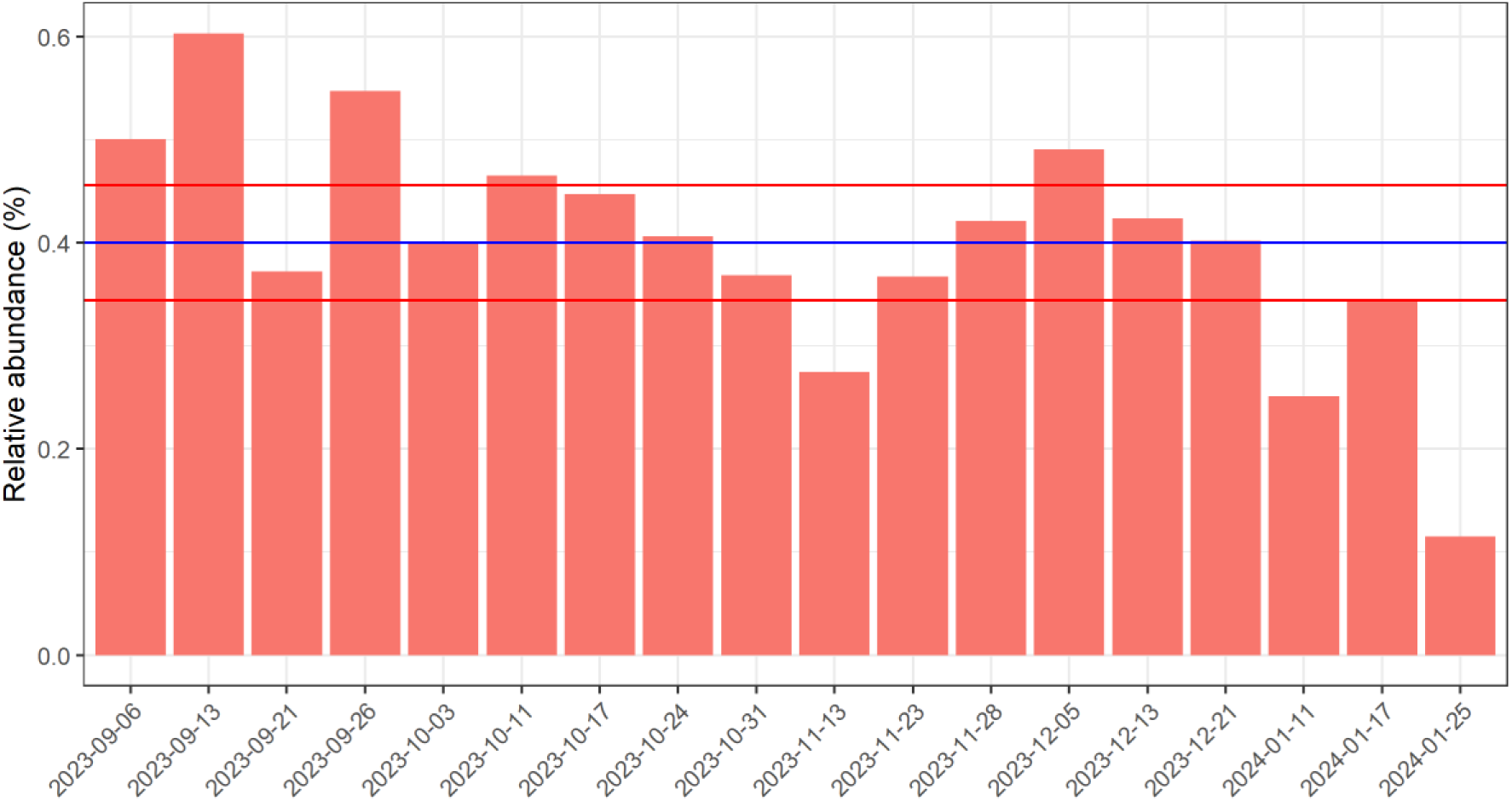
Weekly monitoring of *Escherichia coli* reads in Quebec City raw sewage water samples from September 2023 to January 2024. Blue line: average relative abundance, here defined as the percentage of classified reads within a sample. Red lines: 95% confidence interval.

**Figure 3.**
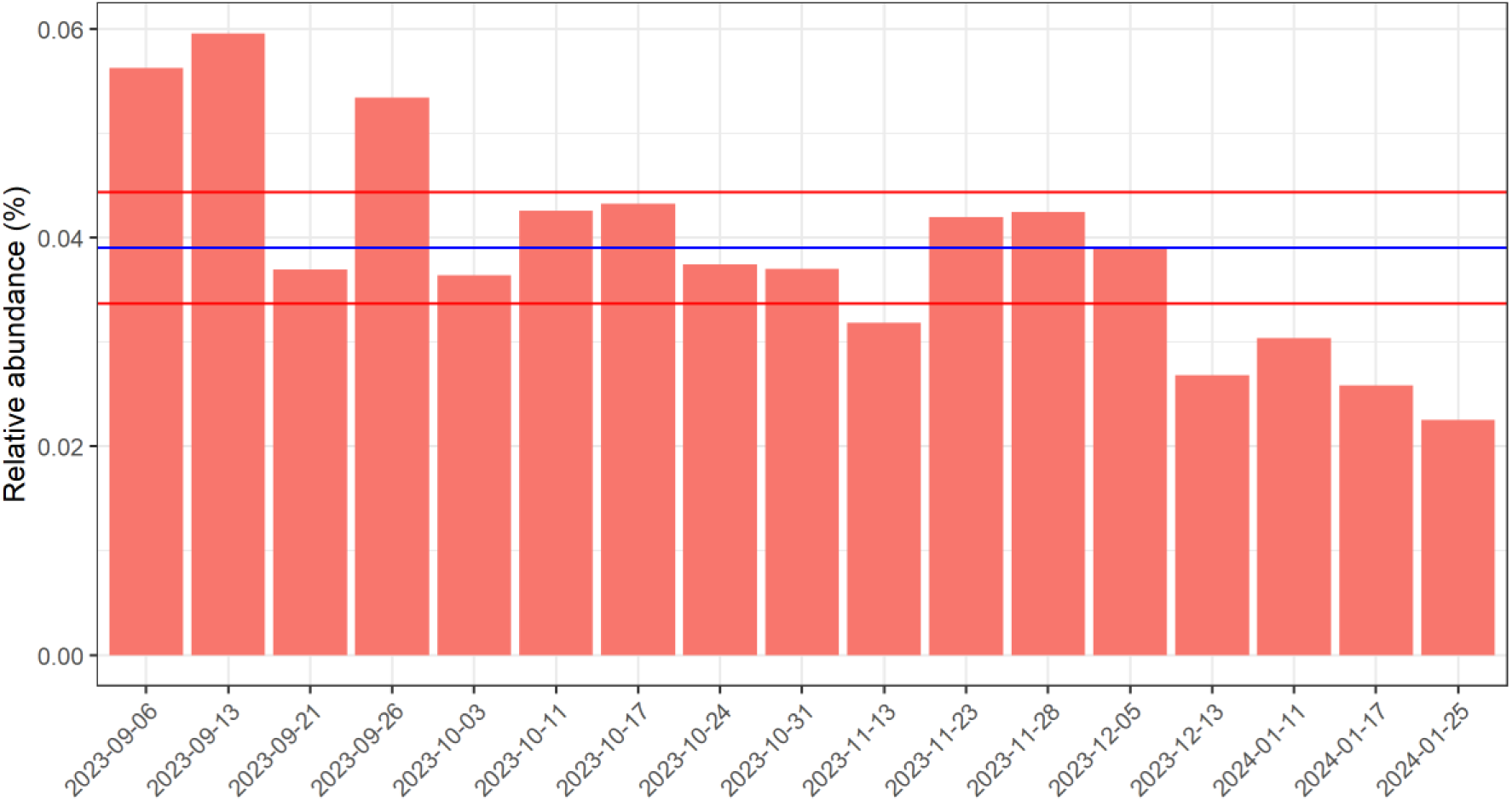
Weekly monitoring of *Salmonella enterica* reads in Quebec City raw sewage water samples from September 2023 to January 2024. Blue line: average relative abundance, here defined as the percentage of classified reads within a sample. Red lines: 95% confidence interval.

### Whole metagenome shotgun assembly

Read-based metagenomic surveillance helped track enterotoxigenic pathogens at the species level. However, non-virulent conspecific strains could also contribute to the abundance peak, given the variable accuracy of read classifiers at the species level [28]. Kraken2 is particularly affected by Nanopore sequence error rates as its algorithm performs exact k-mer matches to classify sequences [11]. Therefore, we attempted to *de novo* assemble the whole metagenome shotgun sequence for the 2023-09-13 sample to further increase taxonomic resolution. However, this assembly did not recover any contigs classified as *E. coli* or *Salmonella* enterica, even though Flye was run with the – meta flag to allow for uneven coverage across contigs [15].

This could be attributable to the very low abundance of those enteric pathogens within the sample (less than 0.5% of all reads). Indeed, reads from the SW2023-09-13 sample revealed that it was largely populated by bacteria (75% of all read counts), among which Pseudomonadota, the phylum comprising STEC and *Salmonella enterica*., accounted for 43% of all bacterial reads (**Figure 4**). The Enterobacteriaceae family represented 5% of all Pseudomonadota within this sample (**Figure 4**). *Escherichia coli* and *Salmonella enterica* reads, regardless of strains, accounted for 11% and 1% respectively of all Enterobacteriaceae reads (**Figure 4**), thereby illustrating a case where most of the dataset does not represent the genomic target of interest.

**Figure 4.**
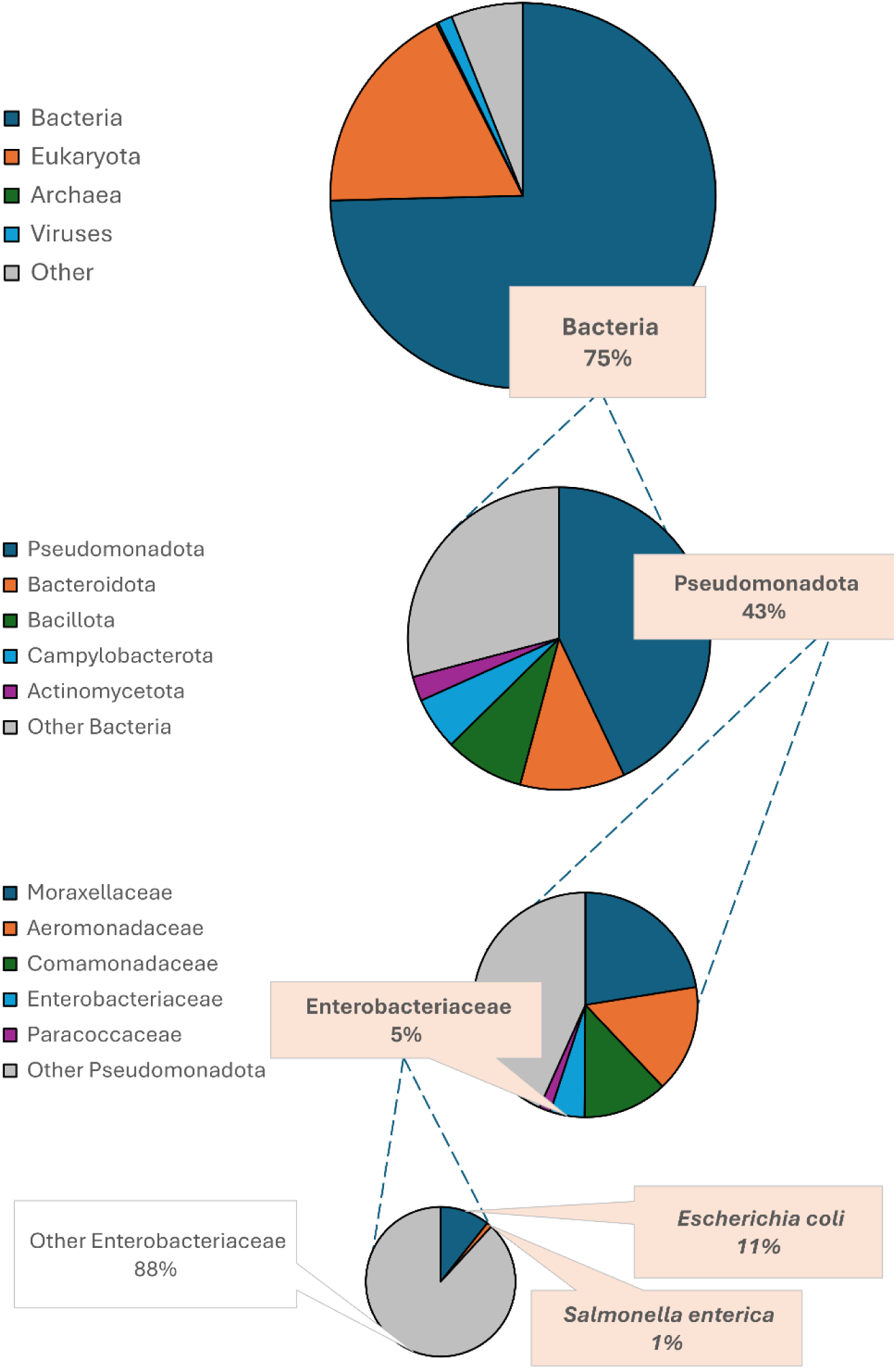
Relative abundance of *Escherichia coli* and *Salmonella enterica* reads from sample SW2023-09-13, expressed as percent read counts within each taxonomic rank. Both species account for 12% of reads classified as Enterobacteriaceae, themselves representing 5% of all read counts within the phylum Pseudomonadota. This phylum itself accounts for 43% of all read counts assigned to Bacteria, themselves comprising 75% of all reads counts within this sample.

We then proceeded to assemble only reads that were assigned as *E. coli* or *S. enterica* using the same *de novo* assembly approach. This approach seemed viable as there were 101,880 reads (935,895,550 bp) assigned to *E. coli*, and 10,574 reads (87,590,049 bp) assigned to *S. enterica.* These subassemblies would have had an expected coverage of 187x and 17.5x coverage assuming 5 Mb genome size. However, these subassemblies were suboptimal for both species. The *E. coli* subassembly had 1,016 contigs totalling 25.4 Mbp with 27x coverage whereas the *S. enterica* subassembly yielded 166 contigs totalling 3,72 Mbp with 9x coverage (less than the expected genome size of 5 Mb for *S. enterica).* Given that SW2023-09-13 was the time point linked with an abundance peak for both species for the whole time series, we do not expect *de novo* subassemblies to perform better on other samples.

Instead of using *de novo* assembly, we attempted to reconstruct genome sequences by reference-guided assemblies using type genomes: O157:H7 strain Sakai for STEC, strain LT2 for enteropathogenic non-typhoid *Salmonella* (ENTS), by using reads from the 2023-09-13 abundance peak as input.

### Reference-guided genome reconstruction

Given the large size of this read set (125 gigabases) and that *E. coli and S. enterica* reads account for 0.4% and 0.04% respectively (**Table 1**), we obtained an incomplete assembly with abnormal feature counts with respect to their reference (**Table 2**). However, when only considering reads assigned to these species (**Table 1**), we obtained sufficient coverage (87.9x and 6.5x) to re-attempt reference-based assemblies. Those assemblies were 95-99% complete (**Table 2** and **Table 3**) and agreed with the main characteristics of their respective reference, thereby indicating that STEC-like and ENTS-like bacteria were contributing factors of this surge.

**Table 1.**
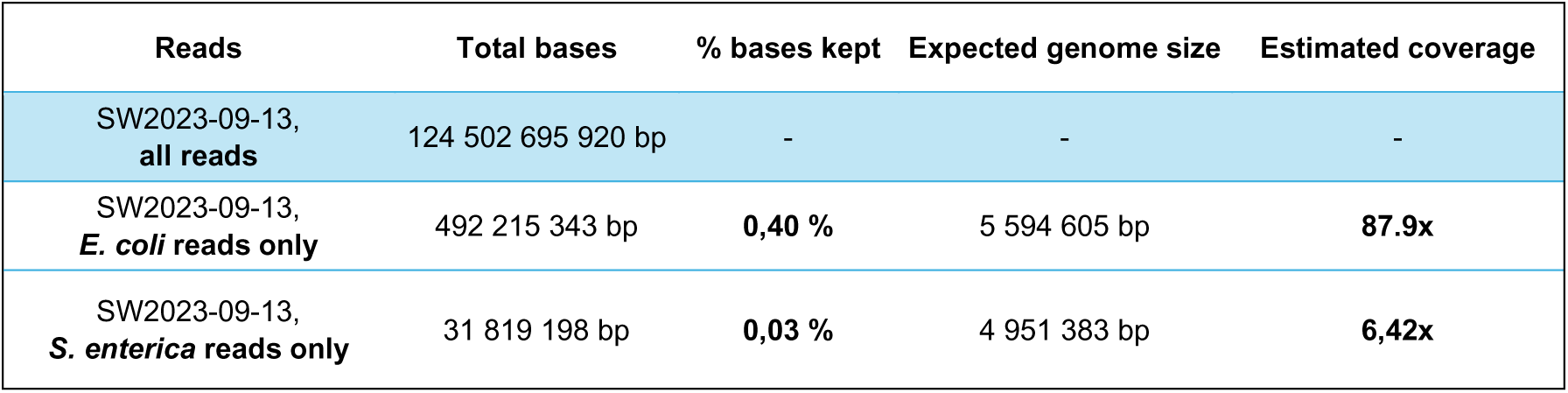
Proportions of reads classified as STEC and *Salmonella enterica* in a predicted abundance peak common to both pathogens of interest (SW2023-09-13).

| Reads | Total bases | % bases kept | Expected genome size | Estimated coverage |
| --- | --- | --- | --- | --- |
| SW2023-09-13,<br>all reads | 124 502 695 920 bp | - | - | - |
| SW2023-09-13,<br>*E. coli* reads only | 492 215 343 bp | 0,40 % | 5 594 605 bp | 87.9x |
| SW2023-09-13,<br>*S. enterica* reads only | 31 819 198 bp | 0,03 % | 4 951 383 bp | 6,42x |

**Table 2.**
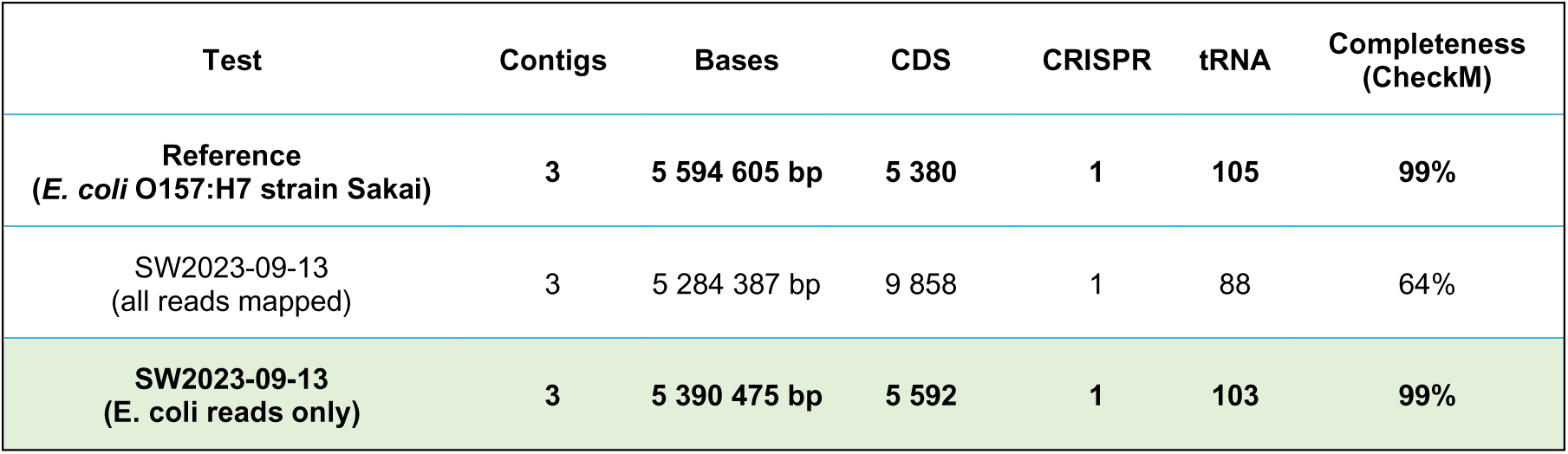
Assembly metrics and features throughout reference-guided genome reconstruction of STEC using nanopore reads from the 2023-09-13 data set.

| Test | Contigs | Bases | CDS | CRISPR | tRNA | Completeness (CheckM) |
| --- | --- | --- | --- | --- | --- | --- |
| Reference<br>(*E. coli* O157:H7 strain Sakai) | 3 | 5 594 605 bp | 5 380 | 1 | 105 | 99% |
| SW2023-09-13<br>(all reads mapped) | 3 | 5 284 387 bp | 9 858 | 1 | 88 | 64% |
| SW2023-09-13<br>(*E. coli* reads only) | 3 | 5 390 475 bp | 5 592 | 1 | 103 | 99% |

**Table 3.** Assembly metrics and features throughout reference-guided genome reconstruction of *S. enterica subsp. enterica* using nanopore reads from the 2023-09-13 data set.

| Test | Contigs | Bases | CDS | CRISPR | tRNA | Completeness (CheckM) |
| --- | --- | --- | --- | --- | --- | --- |
| Reference<br>(*S. enterica* subsp. *enterica*<br>strain LT2) | 2 | 4 951 383 bp | 4 627 | 0 | 86 | 100 % |
| SW2023-09-13<br>(all reads mapped) | 2 | 4 535 144 bp | 8 942 | 0 | 78 | 67 % |
| SW2023-09-13<br>(*S. enterica* reads only) | 2 | 4 930 755 bp | 5 130 | 0 | 79 | 95 % |

Moreover, classical MLST was performed to assess the possibility of typing reference-based assemblies (**Table 4**). In both cases, when all reads from the sample are used to map against the reference, less than 2 alleles out of 7 were identified. In binned-read assemblies, all loci were identified for the STEC-like genome, whereas 5 loci out of 7 were detected for the ENTS-like genome. Interestingly, for the ENTS-like, those 5 loci perfectly matched allele types from *S. enterica* subsp. *enterica* strain LT2 (serovar Typhimurium), suggesting high relatedness to other ENTS strains beyond empirical genome comparisons. We do agree, however, that the degree of representativeness and chimerism in genome reconstruction is a key challenge in metagenomics [29] that can impact the meaningfulness of strain typing on MAGs. The extent of this particular caveat will deserve further investigation.

**Table 4.** Multilocus sequence typing (MLST) analysis for STEC-like and ENTS-like reference-based assemblies with metagenomic data from wastewater sample SW2023-09-13).

| Sample | *adk* | *fumC* | *gyrB* | *icd* | *mdh* | *purA* | *recA* |
| --- | --- | --- | --- | --- | --- | --- | --- |
| Reference<br>(*E. coli* O157:H7 strain Sakai) | 12 | 12 | 8 | 12 | 15 | 2 | 2 |
| SW2023-09-13<br>(all reads mapped) |  |  |  |  | new | new |  |
| SW2023-09-13<br>(*E. coli* reads only) | new | new | 22 | new | 7 | new | 2 |

| Sample | *aroC* | *dnaN* | *hemD* | *hisD* | *purE* | *sucA* | *thrA* |
| --- | --- | --- | --- | --- | --- | --- | --- |
| Reference<br>(*S. enterica* subsp. *enterica* strain LT2) | 10 | 7 | 12 | 9 | new | 9 | 2 |
| SW2023-09-13<br>(all reads mapped) |  |  |  |  |  |  |  |
| SW2023-09-13<br>(*S. enterica* reads only) | 10 | 7 | 12 | 9 | new |  |  |

### Virulence factors

Genome maps also indicated full reference coverage for both the STEC-like and ENTS-like assemblies (**Figure 5**). In the case of the 2023-09-13 STEC-like, the Shiga toxin production genes (*stxAB*) were not only covered by the assembly but were also 100% identical to the reference (**Suppl. Figure 1**). As for the ENTS-like assembly, completeness of the two main type three secretion systems (T3SS) encoded by Salmonella pathogenicity islands 1 and 2 (SPI1 and SPI2) was assessed (**Table 5**). In total, 54 out of 58 genes encoding subunits of the molecular nanosyringe complex were detected in the reconstructed *Salmonella* assembly from 2023-09-13 reads, while 23 of the 26 effectors encoded by *S. enterica* subsp. *enterica* strain LT2 were also present, indicating that T3SS-bearing *S. enterica* most likely contributed to the abundance peak detected on September 13^th^, 2023.

**Figure 5.**
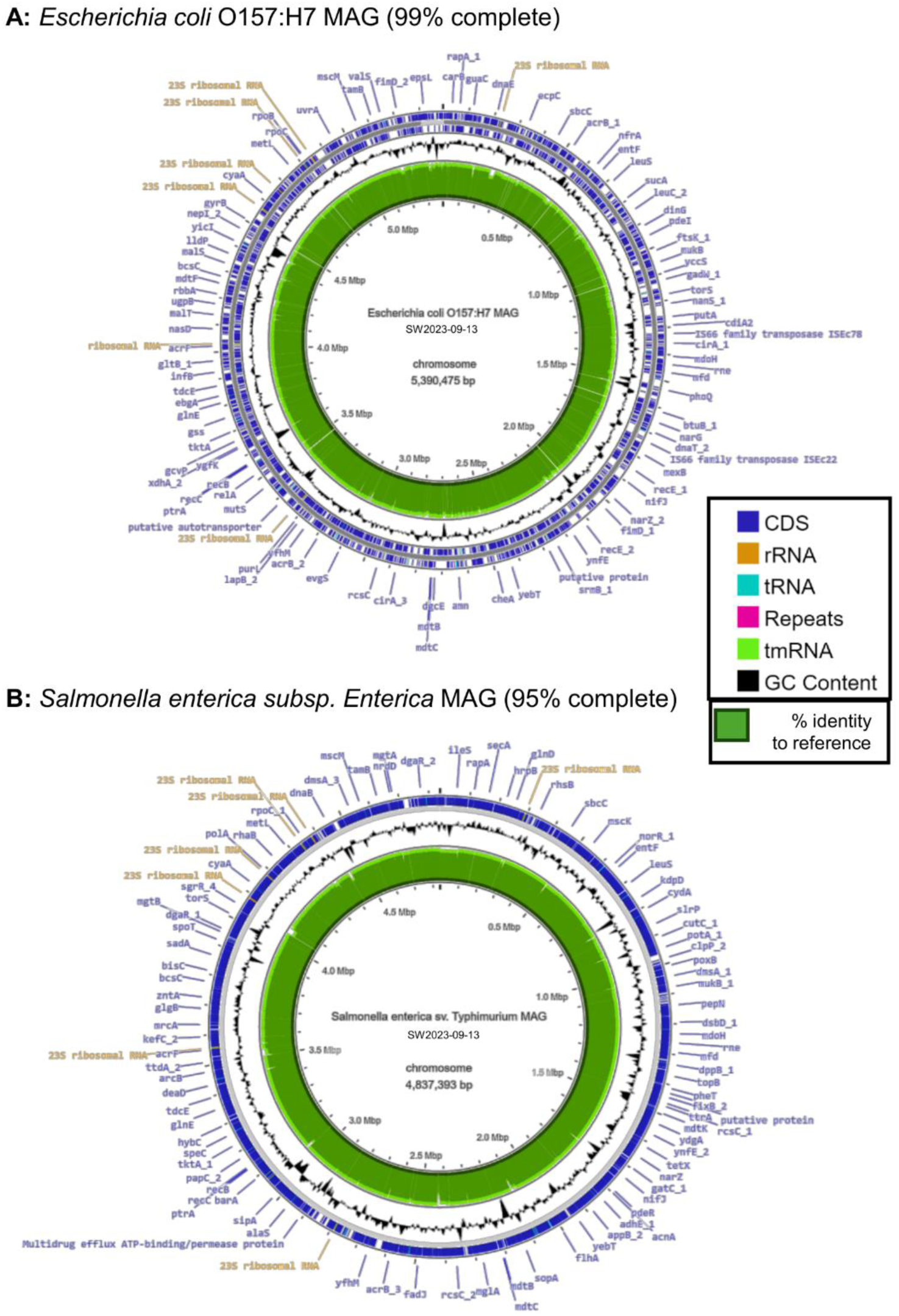
Map of enteric pathogen genomes reconstructed with reads from a pathogen abundance peak (2023-09-13). Panel A: STEC. Panel B: Salmonella enterica serovar Typhimurium. Outer ring: reconstructed contigs. Inner ring: references (GCF_000008865.2 and GCF_000006945.2 respectively for STEC and nontyphoid *Salmonella enterica*).

**Table 5.**
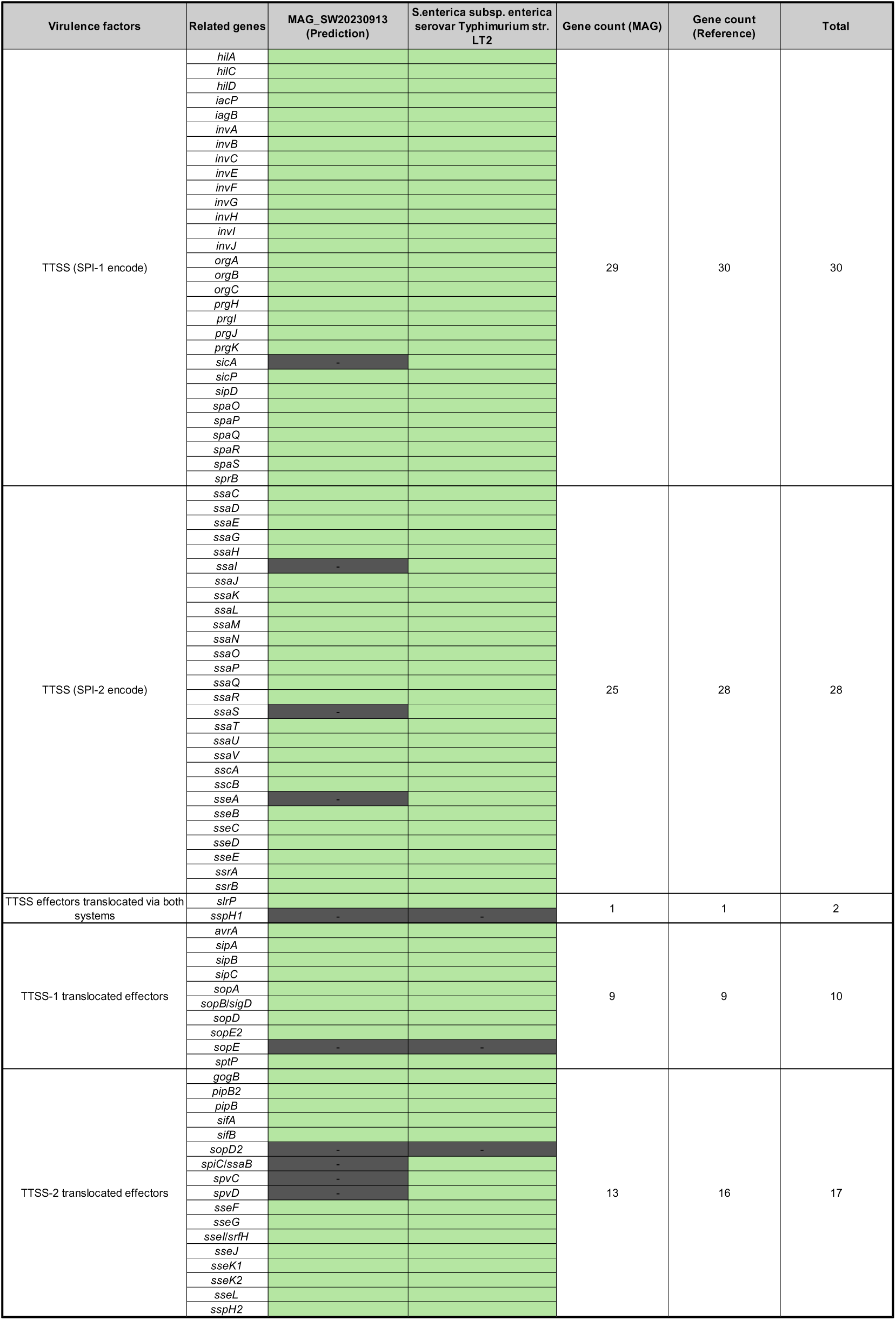
Completeness of the SPI-1 and SPI2-encoded type three secretion system apparatus and effector gene clusters in *Salmonella enterica* reference-based assembly (MAG_SW20230913) and reference (strain LT2).

## Conclusion

High-throughput long-read metagenomic surveillance of Quebec City’s wastewater allowed tracking of two enteric pathogens below 0.1-1% of relative abundance. Shiga toxin producing *Escherichia coli* (STEC) and enteropathogenic non-typhoidal *Salmonella enterica* (ENTS) reached maximum abundance on September 13^th^, 2023, coinciding with initial cases leading to food recalls by Canadian health authorities. Binned-read reference-based assembly further confirmed the presence of STEC and ENTS in the wastewater influent in a way that read taxonomic assignment, or *de novo* metagenome shotgun assembly could not do single-handedly. We therefore suggest that continuous read-based monitoring, coupled with binned-read reference-based assembly for further validation, could become part of a nationwide surveillance pipeline for health authorities.

## Data Availability

Raw sequence reads and genome sequences from this study have been deposited in NCBI databases under the following BioProject primary accession code: PRJNA1134722.

https://www.ncbi.nlm.nih.gov/bioproject/?term=PRJNA1134722

## Ethics approval and consent to participate

Neither human nor animal subjects were part of this study. Wastewater influent sampling for metagenomic analysis has been conducted with the authorization and technical support from Quebec City’s Wastewater Treatment Service (East Station, Quebec, QC, Canada).

## Consent for publication

Not applicable (this study does not contain any individual person’s data in any form).

## Authors’ contributions

J.G. performed bioinformatics analyses and wrote the main manuscript text, figures and tables. S.M. and I.K.I. extracted and sequenced DNA from wastewater samples, discussed results. R.C.L. provided raw wastewater samples and supervised this project alongside S.M., I.K.I., B.B., C.L. and L.G. All authors reviewed the manuscript.

## Acknowledgements

The authors wish to thank Francis Bellavance and Sara Mountir from Managium for project management, Mr. Denis Dufour for technical support at the Quebec wastewater facility treatment plant, Professor Matthew Loose (DeepSeq, University of Liverpool) for training team members and for his support and encouragement in using the Oxford Nanopore dynamic sequencing technology. We also thank Quebec City’s Wastewater Treatment Service for allowing us to take weekly samples.

## References

1. Diamond MB, Keshaviah A, Bento AI, Conroy-Ben O, Driver EM, Ensor KB, et al. Wastewater surveillance of pathogens can inform public health responses. Nat Med. 2022;28:1992–5.

2. O’Neill J. Tackling drug-resistant infections globally: final report and recommendations. Report. Government of the United Kingdom; 2016.

3. Antimicrobial Resistance Collaborators. Global burden of bacterial antimicrobial resistance in 2019: a systematic analysis. Lancet. 2022;399:629–55.

4. Callahan BJ, Wong J, Heiner C, Oh S, Theriot CM, Gulati AS, et al. High-throughput amplicon sequencing of the full-length 16S rRNA gene with single-nucleotide resolution. Nucleic Acids Res. 2019;47:e103.

5. Johnson JS, Spakowicz DJ, Hong B-Y, Petersen LM, Demkowicz P, Chen L, et al. Evaluation of 16S rRNA gene sequencing for species and strain-level microbiome analysis. Nat Commun. 2019;10:5029.

6. Matchado MS, Rühlemann M, Reitmeier S, Kacprowski T, Frost F, Haller D, et al. On the limits of 16S rRNA gene-based metagenome prediction and functional profiling. Microbial Genomics. 2024;10:001203.

7. Joseph TA, Pe’er I. An Introduction to Whole-Metagenome Shotgun Sequencing Studies. Methods Mol Biol. 2021;2243:107–22.

8. Wu Y-W, Simmons BA, Singer SW. MaxBin 2.0: an automated binning algorithm to recover genomes from multiple metagenomic datasets. Bioinformatics. 2016;32:605–7.

9. Kang DD, Li F, Kirton E, Thomas A, Egan R, An H, et al. MetaBAT 2: an adaptive binning algorithm for robust and efficient genome reconstruction from metagenome assemblies. PeerJ. 2019;7:e7359.

10. Vosloo S, Huo L, Anderson CL, Dai Z, Sevillano M, Pinto A. Evaluating de Novo Assembly and Binning Strategies for Time Series Drinking Water Metagenomes. Microbiol Spectr. 9:e01434–21.

11. Wood DE, Lu J, Langmead B. Improved metagenomic analysis with Kraken 2. Genome Biology. 2019;20:257.

12. R Core Team. R: A Language and Environment for Statistical Computing. Vienna, Austria: R Foundation for Statistical Computing; 2019.

13. RStudio Team. RStudio: Integrated Development Environment for R. Boston, MA: RStudio, Inc.; 2016.

14. Bonenfant Q, Noé L, Touzet H. Porechop_ABI: discovering unknown adapters in Oxford Nanopore Technology sequencing reads for downstream trimming. Bioinformatics Advances. 2023;3:vbac085.

15. Kolmogorov M, Yuan J, Lin Y, Pevzner PA. Assembly of long, error-prone reads using repeat graphs. Nat Biotechnol. 2019;37:540–6.

16. Li H. Minimap2: pairwise alignment for nucleotide sequences. Bioinformatics. 2018;34:3094–100.

17. Vaser R, Sović I, Nagarajan N, Šikić M. Fast and accurate de novo genome assembly from long uncorrected reads. Genome Res. 2017;27:737–46.

18. Parks DH, Imelfort M, Skennerton CT, Hugenholtz P, Tyson GW. CheckM: assessing the quality of microbial genomes recovered from isolates, single cells, and metagenomes. Genome Res. 2015;25:1043–55.

19. Grant JR, Enns E, Marinier E, Mandal A, Herman EK, Chen C, et al. Proksee: in-depth characterization and visualization of bacterial genomes. Nucleic Acids Research. 2023;51:W484–92.

20. Biguenet A, Bordy A, Atchon A, Hocquet D, Valot B. Introduction and benchmarking of pyMLST: open-source software for assessing bacterial clonality using core genome MLST. Microb Genom. 2023;9:001126.

21. Seemann T. Prokka: rapid prokaryotic genome annotation. Bioinformatics. 2014;30:2068–9.

22. Liu B, Zheng D, Jin Q, Chen L, Yang J. VFDB 2019: a comparative pathogenomic platform with an interactive web interface. Nucleic Acids Res. 2019;47:D687–92.

23. Dawson DE, Keung JH, Napoles MG, Vella MR, Chen S, Sanderson MW, et al. Investigating behavioral drivers of seasonal Shiga-Toxigenic Escherichia Coli (STEC) patterns in grazing cattle using an agent-based model. PLoS One. 2018;13:e0205418.

24. Nastasijevic I, Schmidt JW, Boskovic M, Glisic M, Kalchayanand N, Shackelford SD, et al. Seasonal Prevalence of Shiga Toxin-Producing Escherichia coli on Pork Carcasses for Three Steps of the Harvest Process at Two Commercial Processing Plants in the United States. Appl Environ Microbiol. 2020;87:e01711–20.

25. Flores Monter YM, Chaves A, Arellano-Reynoso B, López-Pérez AM, Suzán-Azpiri H, Suzán G. Edaphoclimatic seasonal trends and variations of the Salmonella spp. infection in Northwestern Mexico. Infect Dis Model. 2021;6:805–19.

26. Public Health Agency of Canada. Public Health Notice: Outbreak of Salmonella infections linked to Malichita and Rudy brand cantaloupes. 2023. https://www.canada.ca/en/public-health/services/public-health-notices/2023/outbreak-salmonella-infections-malichita-cantaloupes.html. Accessed 9 Jul 2024.

27. Public Health Agency of Canada. Public Health Notice: Outbreak of extensively drug-resistant Salmonella infections linked to raw pet food and contact with cattle. 2024. https://www.canada.ca/en/public-health/services/public-health-notices/2023/outbreak-salmonella-infections-under-investigation.html. Accessed 9 Jul 2024.

28. Van Uffelen A, Posadas A, Roosens NHC, Marchal K, De Keersmaecker SCJ, Vanneste K. Benchmarking bacterial taxonomic classification using nanopore metagenomics data of several mock communities. Sci Data. 2024;11:864.

29. Chang T, Gavelis GS, Brown JM, Stepanauskas R. Genomic representativeness and chimerism in large collections of SAGs and MAGs of marine prokaryoplankton. Microbiome. 2024;12:126.

