## Supplementary material for "Nanopore-based pathogen surveillance allows complete metagenome-assembled genome reconstruction of low-abundance enteric pathogens in wastewater samples": Suppl.

**Supplementary information**

Jeff Gauthier[1], Sima Mohammadi[1] , Irena Kukavica-Ibrulj[1], Brian Boyle[1], Chrystal Landgraff[2], Lawrence Goodridge[3], Roger C. Levesque[1]

1. Département de microbiologie-infectiologie et d'immunologie, Institut de Biologie Intégrative et des Systèmes, Université Laval, Quebec, Canada
2. National Microbiology Laboratory, Public Health Agency of Canada, Winnipeg, MB, Canada
3. Food Science Department, University of Guelph, Guelph, ON, Canada.

### Supplementary Figures

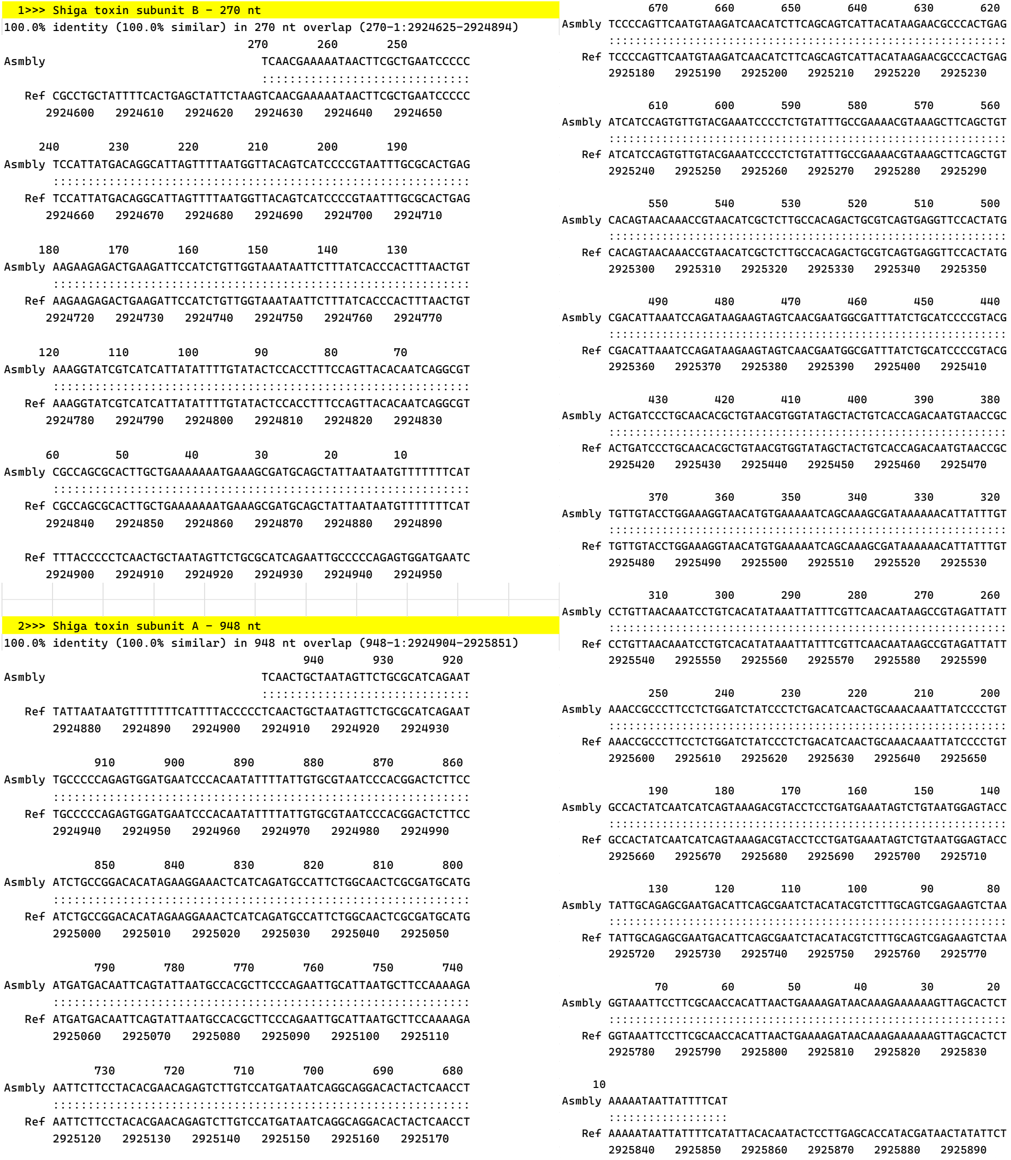


**Suppl. Figure 1.** Pairwise alignment of the Shiga toxin subunit A and B genes between the STEC-like assembly from 2023-09-13 and reference *E. coli* O157:H7 strain Sakai. Assembly: coding DNA sequences for *stxAB* predicted ab initio by PROKKA 1.14.3. Ref: whole genome assembly of *E. coli* O157:H7 strain Sakai.

### Supplementary Tables

**Suppl. Table 1.** Overall species abundance, estimated with Kraken2 by percent read counts, throughout Quebec City wastewater samples sequenced from September 2023 to January 2024.

| Species assignments | Rank | N samples present | % of read counts | 95% confidence interval |
|---|---|---|---|---|
| ***Moraxella osloensis*** | 1 | 18 | 7.12 | 2.02 |
| ***Acinetobacter johnsonii*** | 2 | 18 | 4.43 | 0.46 |
| ***Arcobacter suis*** | 3 | 18 | 2.97 | 0.91 |
| ***Aeromonas media*** | 4 | 18 | 2.03 | 0.29 |
| ***Aliarcobacter cryaerophilus*** | 5 | 18 | 1.96 | 0.35 |
| ***Homo sapiens*** | 6 | 18 | 1.56 | 0.62 |
| ***Aeromonas caviae*** | 7 | 18 | 1.50 | 0.30 |
| ***Acinetobacter sp. TTH0-4*** | 8 | 18 | 1.50 | 0.72 |
| ***Lactococcus raffinolactis*** | 9 | 18 | 1.49 | 0.56 |
| ***Tolumonas auensis*** | 10 | 18 | 1.44 | 0.36 |
| **…** | … | … | … | … |
| ***Escherichia coli*** | 32 | 18 | 0.40 | 0.06 |
| ***Salmonella enterica*** | 245 | 17 | 0.04 | 0.01 |
| ***[…]*** | […] | […] | […] | […] |
| ***TOTAL*** | **2,796** | - | - | - |
